# Nurses’ burnout and associated risk factors during the COVID-19 pandemic: a systematic review and meta-analysis

**DOI:** 10.1101/2020.11.24.20237750

**Authors:** Petros Galanis, Irene Vraka, Despoina Fragkou, Angeliki Bilali, Daphne Kaitelidou

**Author notes:** Corresponding author: Petros Galanis, PhD, Faculty of Nursing, Center for Health Services Management and Evaluation, National and Kapodistrian University of Athens, 123 Papadiamantopoulou street, GR-11527, Athens, Greece. **Author contributions** P.G, I.V. and D.K. were responsible for the conception and design of the study. P.G, I.V., D.F., and A.B. were responsible for the acquisition, analysis and interpretation of data. All the authors drafted the article or revised it critically for important intellectual content, and provided final approval of the version to be submitted.

## Abstract

**Background:** During the COVID-19 pandemic, physical and mental health of the nurses is greatly challenged since they work under unprecedented pressure and they are more vulnerable to the harmful effects of the disease.

**Aim:** To examine the impact of the COVID-19 pandemic on nurses’ burnout and to identify associated risk factors.

**Methods:** We followed the Preferred Reporting Items for Systematic Reviews and Meta-Analysis guidelines for this systematic review and meta-analysis. PubMed, Scopus, ProQuest and pre-print services (medRχiv and PsyArXiv) were searched from January 1, 2020 to November 15, 2020 and we removed duplicates. We applied a random effect model to estimate pooled effects since the heterogeneity between results was very high.

**Findings:** Fourteen studies, including 17,390 nurses met the inclusion criteria. Five standardized and valid questionnaires were used to measure burnout among nurses; Maslach Burnout Inventory, Copenhagen Burnout Inventory, Professional Quality of Life Scale version 5, Mini-Z, and Spanish Burnout Inventory. The overall prevalence of emotional exhaustion was 34.1% (95% confidence interval [CI]: 22.5-46.6%), of depersonalization was 12.6% (95% CI: 6.9-19.7%), and of lack of personal accomplishment was 15.2% (95% CI: 1.4-39.8%). The following factors were associated with increased nurses’ burnout: younger age, higher educational level, higher degree, decreased social support, having a relative/friend diagnosed with COVID-19, low family and colleagues readiness to cope with COVID-19 outbreak, increased perceived threat of Covid-19, longer working time in quarantine areas, working in a high-risk environment (a COVID-19 designated hospital, a COVID-19 unit, etc.), working in hospitals with inadequate and insufficient material and human resources, decreased working safety while caring for COVID-19 patients, increased workload, decreased self-confidence in self-protection, and lower levels of specialized training regarding COVID-19, job experience, and self-confidence in caring for COVID-19.

**Conclusion:** Nurses experience high levels of burnout during the COVID-19 pandemic, while several sociodemographic, social and occupational factors affect this burnout. Several interventions need to be implemented to mitigate mental health impact of the COVID-19 pandemic on nurses, e.g. screening for mental health illness and early supportive interventions for high-risk nurses, immediate access to mental health care services, social support to reduce feelings of isolation, sufficient personal protective equipment for all nurses to provide security etc. Governments, health care organizations and policy makers should act in this direction to prepare health care systems, individuals and nurses for a better response against the COVID-19 pandemic.

## Introduction

In December 2019, the severe acute respiratory syndrome coronavirus 2 (SARS-CoV-2) and related disease (coronavirus disease 2019, COVID-19) emerged from the Wuhan, capital of Hubei province, China^1^. The spread of SARS-CoV-2 infection is much broader than other recent epidemic infections (SARS, MERS)^2^ resulting on 58,712,326 cases globally and 1,388,528 deaths by November 24, 2020^3^.

Health care workers (HCWs) represent a high risk group for SARS-CoV-2 infection since the proportion of HCWs who are positive among all COVID-19 patients is 10.1%^4^. Also, seroprevalence of SARS-CoV-2 antibodies among HCWs^5^ is higher than general population^6^ worldwide (8.7% vs. 5.3%) indicating the higher probability of HCWs to be infected. SARS-CoV-2 can be transmitted by symptomatic, pre-symptomatic, or asymptomatic infected individuals and the proportion of asymptomatic infection among COVID-19 carriers seems to be substantial^7^ putting healthcare systems under extreme pressure and posing a significant threat to public health.

During the COVID-19 pandemic, physical and mental health of the HCWs is greatly challenged since they work under unprecedented pressure and they are more vulnerable to the harmful effects of the disease. Several systematic reviews^8–20^ highlights the tremendous impact of COVID-19 pandemic on psychological and mental health of HCWs representing a high risk group for develop a wide range of problems. In particular, the prevalence of anxiety, depression, insomnia, sleep disturbances, burnout, fear, post-traumatic stress disorder, psychological distress is increased among HCWs during the COVID-19 pandemic^8–20^. Several factors increase theses problems among HCWs e.g. direct contact with COVID-19 patients, low social support, working in the nursing profession, fewer years of job experience, female gender, work overload, working in a high-risk environment, and absence of specialized training^12,15–19^.

Nurses play an instrumental role to the health systems response to COVID-19 pandemic since they are the frontline health care workers directly involved in the treatment and care of patients. Nurses are under extreme and persistent psychological pressure since they are particularly exposed to the threat of SARS-CoV-2 infection and they become overwhelmed by fear for the safety for their own health, their close family members and their patients. Under these circumstances nurses experience severe psychological and mental problems that could lead to burnout and then to lower productivity, errors in clinical settings, and lack of concern in handling patients. Since the second wave of COVID-19 is hitting worldwide, health care facilities with exhausted nurses are the worst scenario to confront the pandemic.

To our knowledge, there is no systematic review to emphasize in nurses and especially in burnout during the COVID-19 pandemic. Thus, given the exponential increase of studies on this research area we aimed to examine the impact of the COVID-19 pandemic on nurses’ burnout and to identify risk factors associated with burnout.

## Methods

### Data sources and strategy

We followed the Preferred Reporting Items for Systematic Reviews and Meta-Analysis (PRISMA) guidelines for this systematic review and meta-analysis^21^. PRISMA checklist is presented in Web Table 1. PubMed, Scopus, ProQuest and pre-print services (medRχiv and PsyArXiv) were searched from January 1, 2020 to November 15, 2020 and we removed duplicates. We used the following strategy: ((nurs* OR “nursing staff” OR “health personnel” OR “healthcare workers” OR “healthcare personnel” OR “health care personnel” OR “health care workers”) AND (COVID-19 OR SARS-COV-2)) AND (burnout). Search in Pubmed, medRχiv and PsyArXiv included title/abstract, while in Scopus and ProQuest included title/abstract/keywords. Also, we searched the full reference lists of all selected articles.

**Table 1.** Main characteristics of the studies included in this systematic review.

| Reference | Location | Females (%) | Age, mean (SD) | Sample size (n) | Study design | Sampling method | Assessment tool | Response rate (%) | Data collection time | Publication in |
| --- | --- | --- | --- | --- | --- | --- | --- | --- | --- | --- |
| Chen et al. 2020<sup>27</sup> | China and Taiwan | 95.6 | 33.1 (7.5) | 12,596 | Cross-sectional | Convenience sampling | MBI | NR | April | Journal |
| Aydin Sayilan et al. 2020<sup>28</sup> | Turkey | 75.3 | 28 (6) | 267 | Cross-sectional | Convenience sampling | MBI | 69.5 | May 10-20 | Journal |
| Buselli et al. 2020<sup>29</sup> | Italy | NR | NR | 133 | Cross-sectional | Convenience sampling | ProQOL-5 | NR | April 1 to May 1 | Journal |
| Chor et al. 2020<sup>30</sup> | Singapore | NR | NR | 210 | Cross-sectional | Convenience sampling | CBI | 55.7 | May | Journal |
| Cortina-Rodríguez et al. 2020<sup>31</sup> | Puerto Rico | NR | NR | 23 | Cross-sectional | Snowball sampling | MBI | NR | April 25 to May 25 | Pre-print service |
| Ferry et al. 2020<sup>32</sup> | United Kingdom | NR | NR | 286 | Cross-sectional | Snowball sampling | CBI | NR | June 17-24 | Pre-print service |
| Hu et al. 2020<sup>33</sup> | China | 87.1 | 31 (6.2) | 2101 | Cross-sectional | Convenience sampling | MBI | 99.6 | February 13-24 | Journal |
| Khasne et al. 2020<sup>34</sup> | India | NR | NR | 198 | Cross-sectional | Convenience sampling | CBI | NR | NR | Journal |
| Jalili et al. 2020<sup>35</sup> | Iran | NR | NR | 300 | Cross-sectional | Convenience sampling | MBI | NR | NR | Pre-print service |
| Manzano-Garcia et al. 2020<sup>36</sup> | Spain | 90 | 42.4 (11.4) | 771 | Cross-sectional | Convenience sampling | SBI | 39 | April 15-30 | Journal |
| Matsuo et al. 2020<sup>37</sup> | Japan | 126 | NR | NR | Cross-sectional | Convenience sampling | MBI | NR | April 6-19 | Journal |
| Prasad et al. 2020<sup>38</sup> | USA | NR | NR | NR | Cross-sectional | Convenience sampling | Mini-Z | NR | April 14-25 | Journal |
| Ruiz-Fernandez et al. 2020<sup>39</sup> | Spain | NR | NR | 398 | Cross-sectional | Convenience sampling | ProQoL-5 | NR | March 30 to April 16 | Journal |
| Zhang et al. 2020<sup>40</sup> | China | 90.7 | 30.3 (5.5) | 107 | Cross-sectional | Convenience sampling | MBI | 97 | March 10-14 | Journal |

CBI: Copenhagen Burnout Inventory; MBI: Maslach Burnout Inventory; NR: not reported; ProQOL-5: Professional Quality of Life Scale version 5; SD: standard deviation; SBI: Spanish Burnout Inventory

### Selection and eligibility criteria

We initially screened title and abstract of the records and then full-text. Two independent authors performed study selection and disagreements were resolved by a third, senior author. We included studies that were published in English, except case reports, qualitative studies, reviews, protocols, editorials, and letters to the Editor. Also, we included studies examining nurses’ burnout and associated risk factors during the COVID-19 pandemic. Moreover, we included studies that used standardized and valid instruments to measure burnout. All types of nurses working in hospitals that treat COVID-19 patients were accepted for inclusion. We excluded studies reporting results in total for HCWs and not separately for nurses.

### Data extraction and quality assessment

We used structured forms to extract data from each study, such as authors, location, gender, age, sample size, study design, sampling method, assessment tool, response rate, data collection time, publication (journal or pre-print service), number of nurses with burnout, scores on burnout scales, factors associated with burnout, and the level of analysis (univariate or multivariable).

We assessed quality of studies included using the Joanna Briggs Institute critical appraisal tools^22^. This scale is composed of 9 items for prevalence studies, of 8 items for cross-sectional studies, and of 11 items for cohort studies. Studies quality can be poor, moderate or good according to scale score. All studies in our review were cross-sectional, where a score of 7-8 points indicates good quality, a score of 4-6 points indicates moderate quality and a score ≤3 indicates poor quality.

One reviewer extracted the data and assessed the quality for all studies and a second reviewer checked this information for validity and completeness.

### Statistical analysis

For each study we extracted the sample size and the number of nurses that experienced burnout according to scores on burnout scales. Then, we calculated the prevalence of burnout and the 95% confidence interval (CI) for each included study. Prevalences were transformed with the Freeman-Tukey Double Arcsine method before pooling^23^. We used the Hedges Q statistic and I^2^ statistics to assess between-studies heterogeneity. A p-value<0.1 for the Hedges Q statistic indicates statistically significant heterogeneity, while I^2^ values higher than 75% indicates high heterogeneity^24^. We applied a random effect model to estimate pooled effects since the heterogeneity between results was very high^24^. A priori, we considered sample size, studies quality, publication type (journal or pre-print service) and the continent that studies were conducted as sources of heterogeneity. Due to the limited variability of these variables and the limited number of studies, we decided to perform meta-regression analysis only with sample size as the independent variable. We performed a leave-one-out sensitivity analysis to determine the influence of each study on the overall effect. We used a funnel plot and the Egger’s test to assess the publication bias with a P-value<0.05 indicating publication bias^25^. We did not perform meta-analysis for the risk factors associated with nurses’ burnout since the data were very limited and highly heterogeneous. Statistical analysis was performed with OpenMeta[Analyst]^26^.

## Results

### Identification and selection of studies

Flowchart of the literature search is presented in Figure 1 according to PRISMA guidelines. Initially, we identified 375 potential records through electronic databases and 245 were left after removing duplicates. After the screening of the titles and abstracts, we removed 191 records and we added 4 more records found by the reference lists scanning. Finally, we included 14 studies^27–40^ in this systematic review that met our inclusion criteria and among them six studies in the meta-analysis that included the appropriate data.

**Figure 1.**
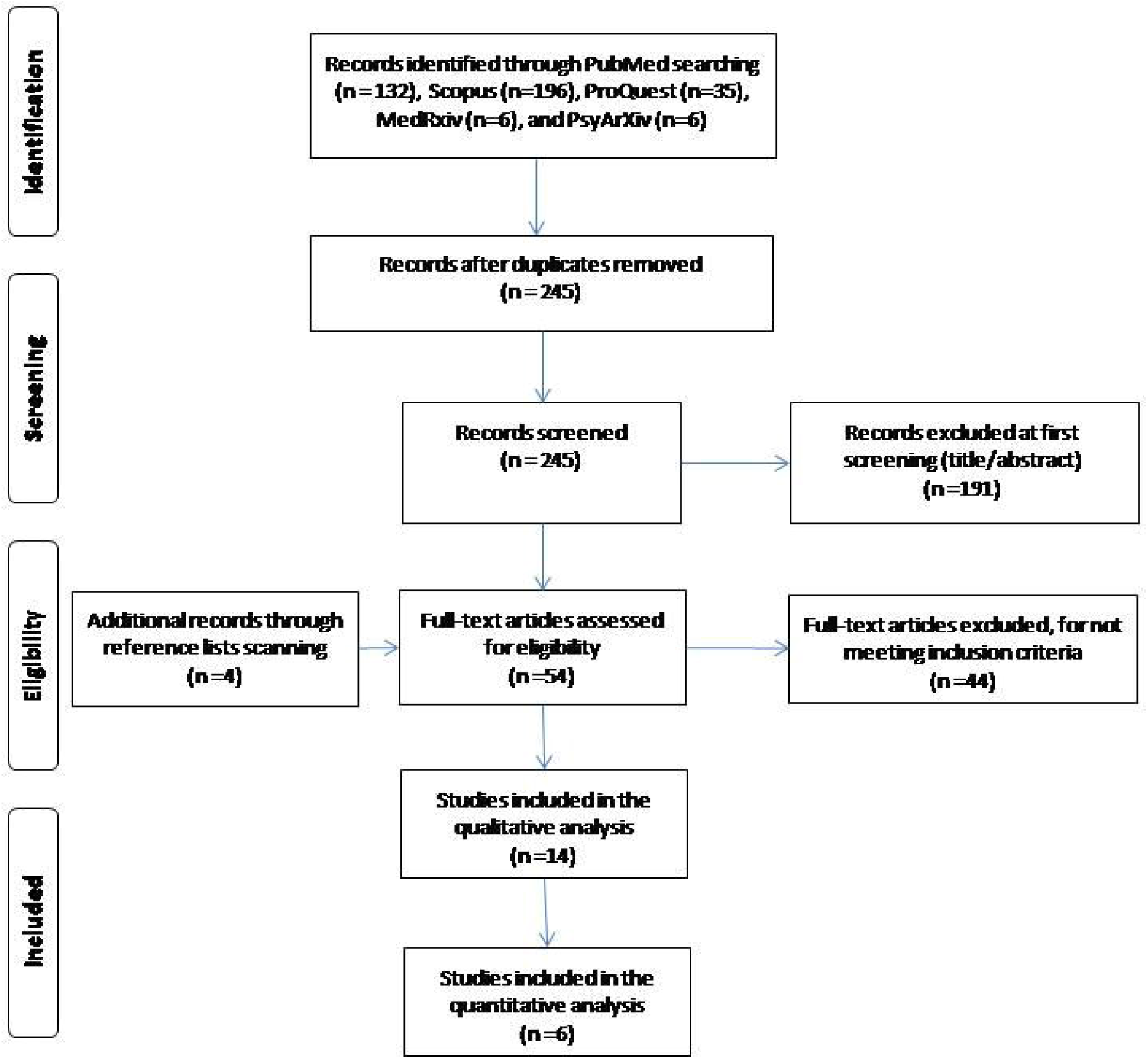
Flowchart of the literature search according to the Preferred Reporting Items for Systematic Reviews and Meta-Analysis.

### Characteristics of the studies

Main characteristics of the 14 studies included in our systematic review are shown in Table 1. Study population included only nurses in five studies,^27,28,33,36,40^ while nine studies^29–32,34,35,37–39^ included HCWs in general. A total of 17,390 nurses were included in this systematic review, while two studies^37,38^ did not report the number of nurses. All studies were cross-sectional, while 12 studies^27–30,33–40^ used a convenience sample method and two studies^31,32^ used a snowball sampling method. Eight studies^27,28,30,33–35,37,40^ was conducted in Asia (China, India, Japan, Turkey, Singapore, Taiwan, and Iran), four studies^29,32,36,39^ in Europe (United Kingdom, Spain, and Italy), and two studies^31,38^ in North America (USA and Puerto Rico). Eleven studies^27– 30,33,34,36–40^ were published in journals and three studies^31,32,35^ in pre-print services. Limited data were available regarding response rate, gender and age.

### Measurement tools for burnout

Five standardized and valid questionnaires were used to measure burnout among nurses. The majority of studies^27,28,31,33,35,37,40^ used the Maslach Burnout Inventory (MBI), three studies^30,32,34^ used the Copenhagen Burnout Inventory (CBI), two studies^29,39^ used the Professional Quality of Life Scale version 5 (ProQOL-5), one study^38^ used the Mini-Z and one study^36^ used the Spanish Burnout Inventory (SBI). The MBI is the most widely used instrument to measure burnout comprising 22 items in three domains; emotional exhaustion, depersonalization and personal accomplishment^41^. Emotional exhaustion score ranges from 0 to 54, depersonalization score ranges from 0 to 30, and personal accomplishment score ranges from 0 to 48. Higher scores on the emotional exhaustion and the depersonalization subscales indicate a higher burnout symptom burden, while lower scores on the personal accomplishment subscale indicates a higher burnout symptom burden. Individuals are classified in low, moderate or high burnout level according to cut-off points for the MBI subscales. The CBI consists of three subscales; personal burnout (six items), work-related burnout (seven items), and client-related burnout (six items)^42^. Subscales score range from 0 to 100 with higher values indicating higher burnout symptom burden. According to the CBI scores, individuals are classified in low, moderate or high burnout level. The ProQOL-5 consists of three subscales; compassion satisfaction (ten items), burnout (ten items), and secondary traumatic stress (ten items)^43^. Higher burnout subscale score indicates increased burnout with individuals classified in low, moderate or high burnout level according to specific cut-off points. The Mini-Z instrument includes ten items and one item is about burnout dividing individuals into two groups; with or without burnout^44^. The SBI consists of three subscales: enthusiasm for the work (five items), psychological exhaustion (four items), and indolence (six items). Total SBI score ranges from 0 to 4 with higher values indicating higher burnout syndrome risk^45^.

### Quality assessment

Quality assessment of cross-sectional studies in this review is shown in Table 2. Quality was moderate in the majority of studies (n=12)^29,29–40^ and good in two studies^27,28^. The most frequent bias is that the studies did not take into account confounding factors and did not multivariable methods to eliminate them.

**Table 2.**
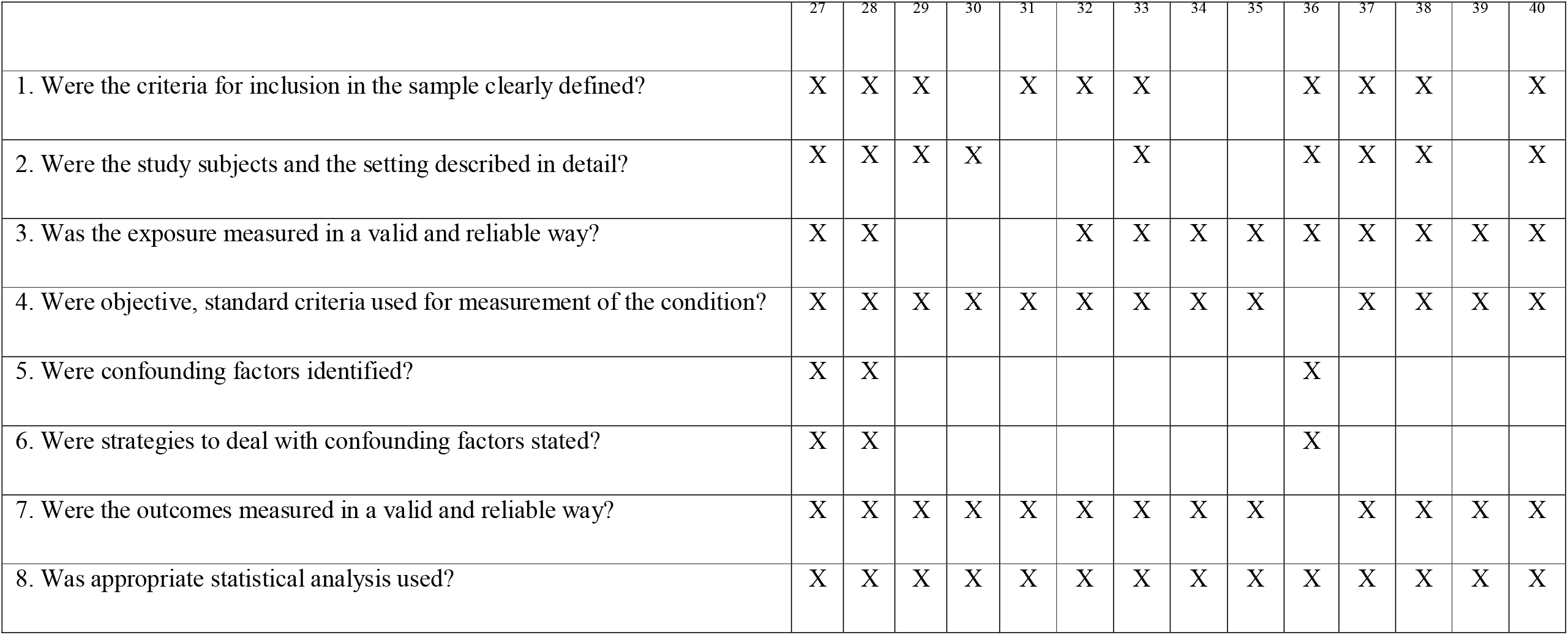
Quality of cross-sectional studies included in this systematic review.

### Meta-analysis

Data regarding burnout scales were highly heterogeneous. Regarding the most frequent tool to measure burnout (MBI), the prevalence of nurses’ burnout in three subscales (emotional exhaustion, depersonalization, and personal accomplishment) was measured in six studies,^27,28,31,33,35,40^ while the mean burnout scores and standard deviations were measured in four studies^27,28,33,40^. Four different studies^29,30,36,39^ measured total mean burnout score with four different instruments, while four other studies^32,34,37,38^ measured the prevalence of total burnout with three different instruments making the synthesis of these results in meta-analysis models counterintuitive. Thus, we decided to include in the meta-analysis the six studies^27,28,31,33,35,40^ that calculated the prevalence of nurses’ burnout in three subscales of the MBI to improve comprehensiveness and clarity.

Descriptive statistics for nurses’ burnout according to the measurement tools used in the studies included in this systematic review and meta-analysis are shown in Table 3. The overall prevalence of emotional exhaustion among nurses according to the MBI was 34.1% (95% CI: 22.5-46.6%) (Figure 2). The prevalence of emotional exhaustion ranged from 5.6% to 69.6% with a very high heterogeneity between results (I^2^=98.9%, p-value for the Hedges Q statistic < 0.001). According to meta-regression analysis, the prevalence was independent of the sample size (p=0.39). A leave-one-out sensitivity analysis showed that removal of studies had an influential effect on the pooled prevalence, which varied between 29.3% (95% CI: 17.9-42.2%), with Cortina-Rodríguez et al.^31^ excluded, and 41.4% (95% CI: 28.2-55.2%), with Zhang et al.^40^ excluded (Web Figure 1). The asymmetrical shape of the funnel plot (Web Figure 2) and p-value<0.05 for Egger’s test implied potential publication bias.

**Table 3.** Descriptive statistics for nurses’ burnout scales according to the measurement tools used in the studies included in this systematic review.

| Reference | Assessment tool | Emotional exhaustion | Depersonalization | Lack of personal accomplishment | Total burnout | Sample size |
| --- | --- | --- | --- | --- | --- | --- |
|  |  | No. of nurses<br>Mean score (SD) | No. of nurses<br>Mean score (SD) | No. of nurses<br>Mean score (SD) | No. of nurses<br>Mean score (SD) |  |
| Chen et al. 2020<sup>27</sup> | MBI | 2709<br>19.1 (10) | 2279<br>5.5 (4.6) | 145<br>19 (8.4) | NR | 12,596 |
| Aydin Sayilan et al. 2020<sup>28</sup> | MBI | 90<br>23.7 (7.9) | 3<br>17.1 (4.6) | 0<br>17.6 (4.1) | NR | 267 |
| Buselli et al. 2020<sup>29</sup> | ProQOL-5 | NR | NR | NR | 19.9 (4.7) | 133 |
| Chor et al. 2020<sup>30</sup> | CBI | NR | NR | NR | 112<br>51.3 (19.6) | 210 |
| Cortina-Rodríguez et al. 2020<sup>31</sup> | MBI | 16<br>32 (NR) | 9<br>9.8 (NR) | 12<br>32.7 (NR) | NR | 23 |
| Ferry et al. 2020<sup>32</sup> | CBI | NR | NR | NR | 245 | 286 |
| Hu et al. 2020<sup>33</sup> | MBI | 835<br>23.4 (13.8) | 556<br>6.8 (7.1) | 771<br>34.8 (10) | NR | 2101 |
| Khasne et al. 2020<sup>34</sup> | CBI | *Personal burnout*<br>101 | *Work-related burnout*<br>76 | *Pandemic-related burnout*<br>96 | NR | 198 |
| Jalili et al. 2020<sup>35</sup> | MBI | 159 | 40 | 1 |  | 300 |
| Manzano-Garcia et al. 2020<sup>36</sup> | SBI | NR | NR | NR | 2.5 (0.3) | 771 |
| Matsuo et al. 2020<sup>37</sup> | MBI | NR | NR | NR | 59 | 126 |
| Prasad et al. 2020<sup>38</sup> | Mini-Z | NR | NR | NR | 83 | 104 |
| Ruiz-Fernandez et al. 2020<sup>39</sup> | ProQoL-5 | NR | NR | NR | 24.3 (5.7) | 398 |
| Zhang et al. 2020<sup>40</sup> | MBI | 6<br>12.3 (7.1) | 2<br>2.1 (2.8) | 52<br>16.5 (8.4) | NR | 107 |

CBI: Copenhagen Burnout Inventory; MBI: Maslach Burnout Inventory; NR: not reported; ProQOL-5: Professional Quality of Life Scale version 5; SD: standard deviation; SBI: Spanish Burnout Inventory

**Figure 2.**
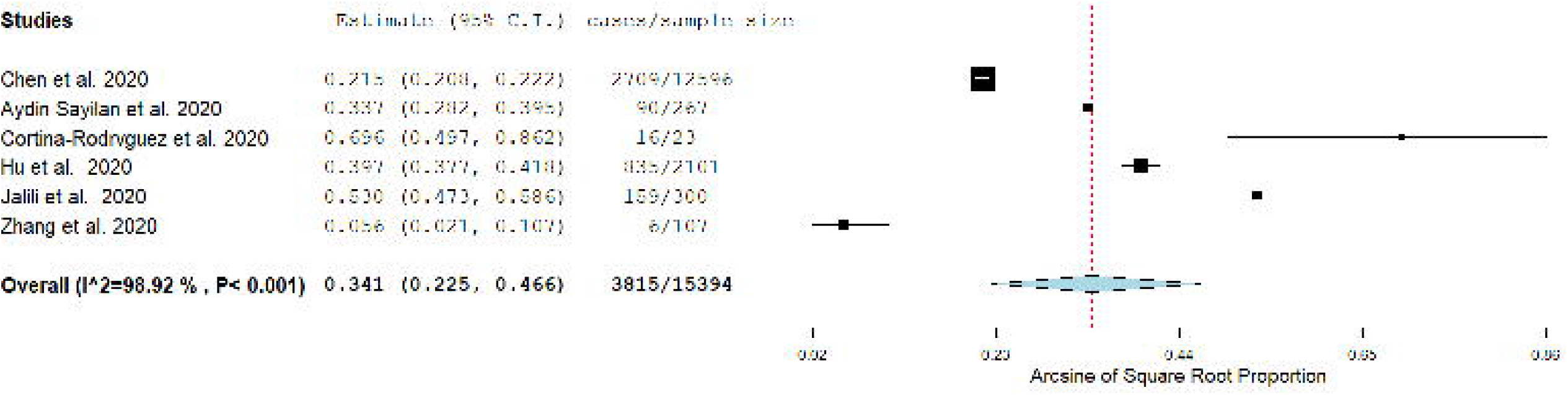
Forest plot of the prevalence of emotional exhaustion among nurses according to the Maslach Burnout Inventory.

Regarding nurses’ depersonalization according to the MBI, the pooled prevalence was 12.6% (95% CI: 6.9-19.7%) (Figure 3), while the sample size did not affect it (p=0.57). The heterogeneity between results was very high (I^2^=98%, p-value for the Hedges Q statistic < 0.001). Sensitivity analysis showed that removal of studies had a slight effect on the prevalence, which varied between 10.3% (95% CI: 5-17.3%), with Cortina-Rodríguez et al.^31^ excluded, and 16.3% (95% CI: 10.7-22.9%), with Aydin Sayilan et al.^28^ excluded (Web Figure 3). Funnel plot (Web Figure 4) and Egger’s test (p-value<0.05) indicated potential publication bias.

**Figure 3.**
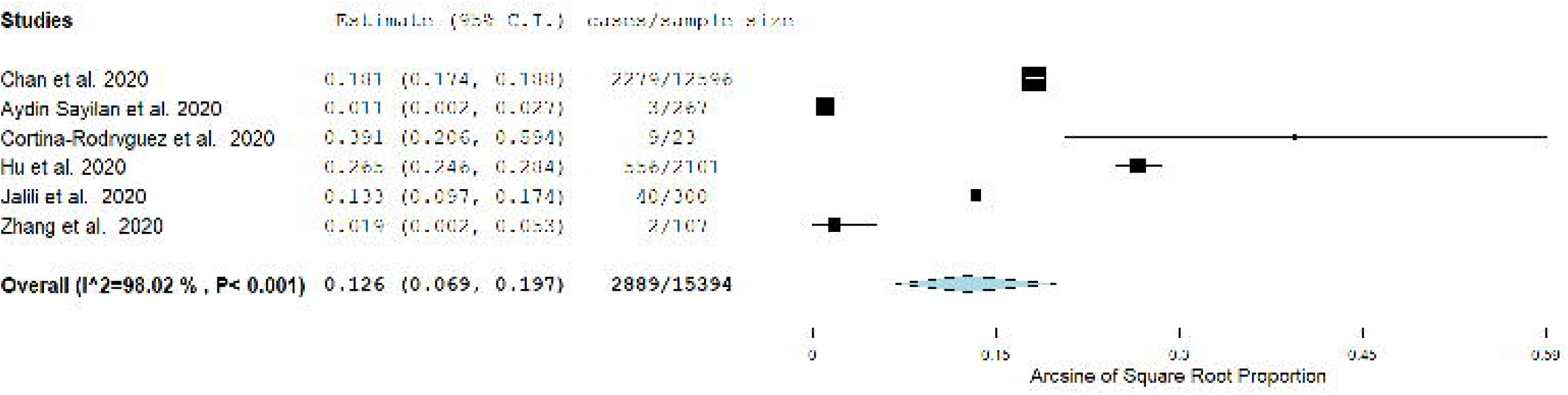
Forest plot of the prevalence of depersonalization among nurses according to the Maslach Burnout Inventory.

The overall prevalence of nurses’ lack of personal accomplishment according to the MBI was 15.2% (95% CI: 1.4-39.8%) (Figure 4). Very high heterogeneity between results was identified (I^2^=99.8%, p-value for the Hedges Q statistic < 0.001). Meta-regression analysis showed that the prevalence was independent of the sample size (p=0.34). According to leave-one-out sensitivity analysis, removal of studies had a moderate effect on the prevalence, which varied between 10.2% (95% CI: 0-34.9%), with Cortina-Rodríguez et al.^31^ excluded, and 20.8% (95% CI: 2.2-51.2%), with Aydin Sayilan et al.^28^ excluded (Web Figure 5). The asymmetrical shape of the funnel plot (Web Figure 6) and p-value<0.05 for Egger’s test implied potential publication bias.

**Figure 4.**
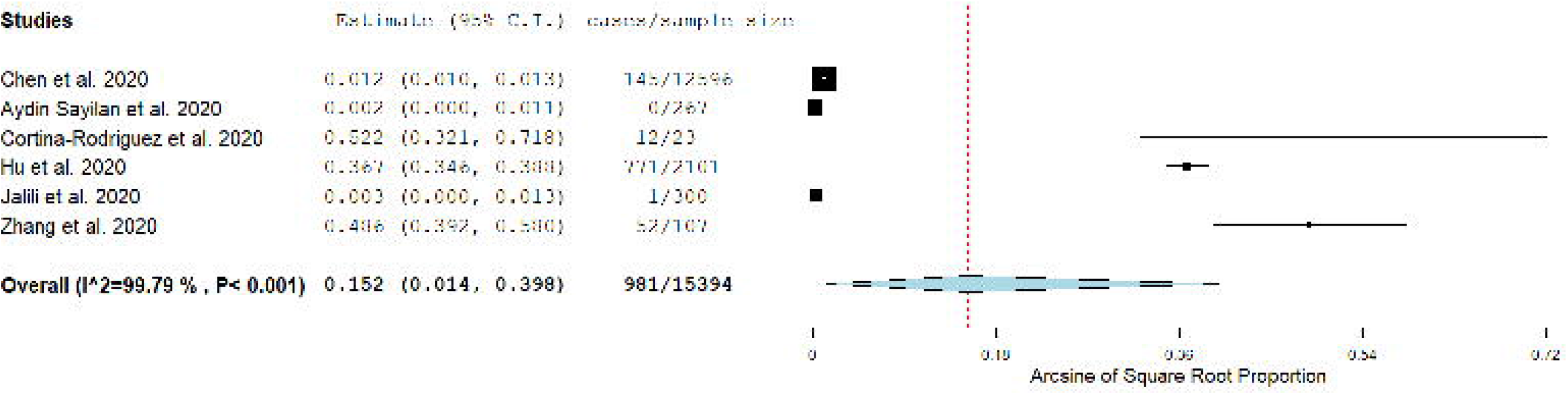
Forest plot of the prevalence of lack of personal accomplishment among nurses according to the Maslach Burnout Inventory.

### Risk factors for burnout

Five studies^27,28,33,36,40^ investigated risk factors for nurses’ burnout during the COVID-19 pandemic and among them three^27,28,36^ used multivariable models to eliminate confounding factors (Table 4).

**Table 4.**
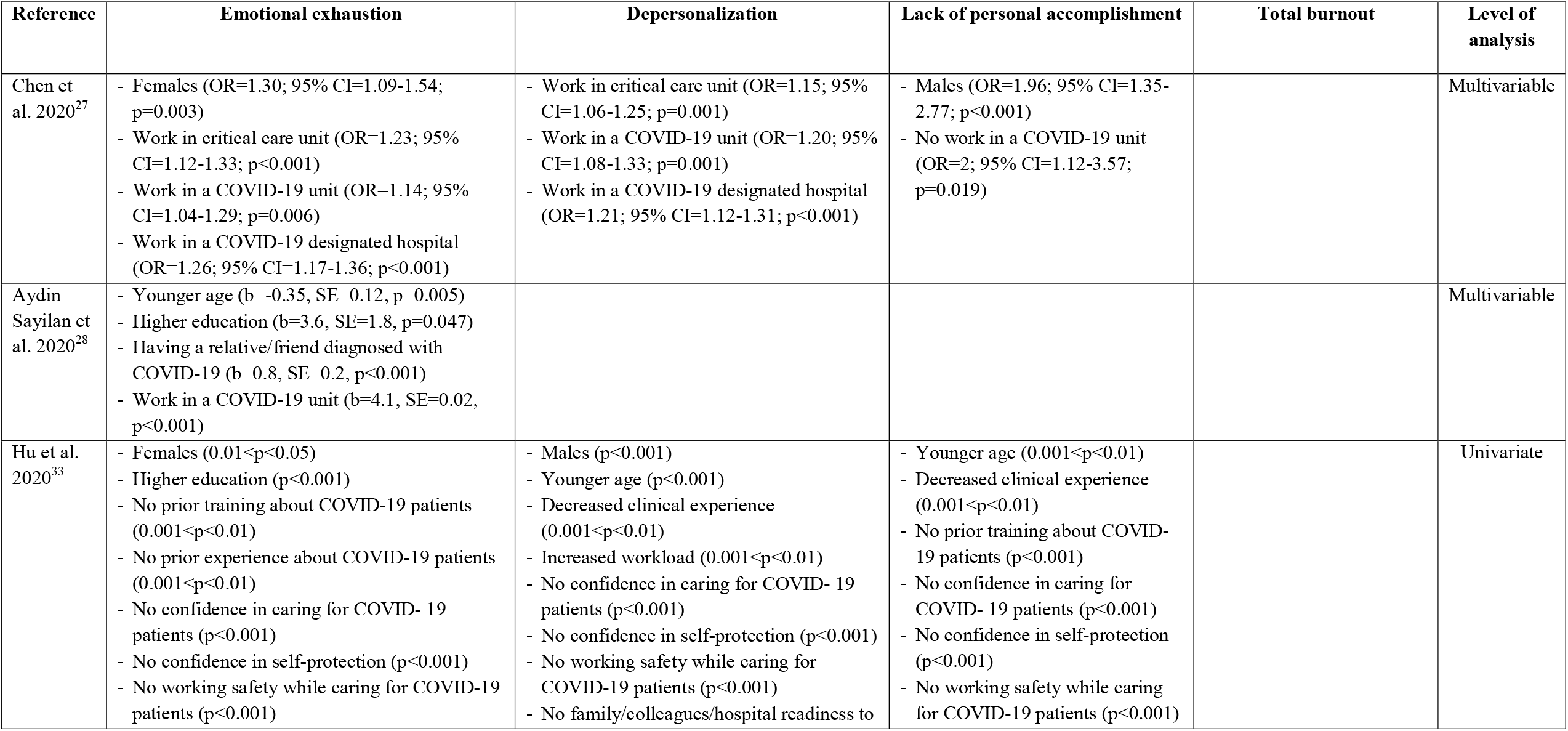
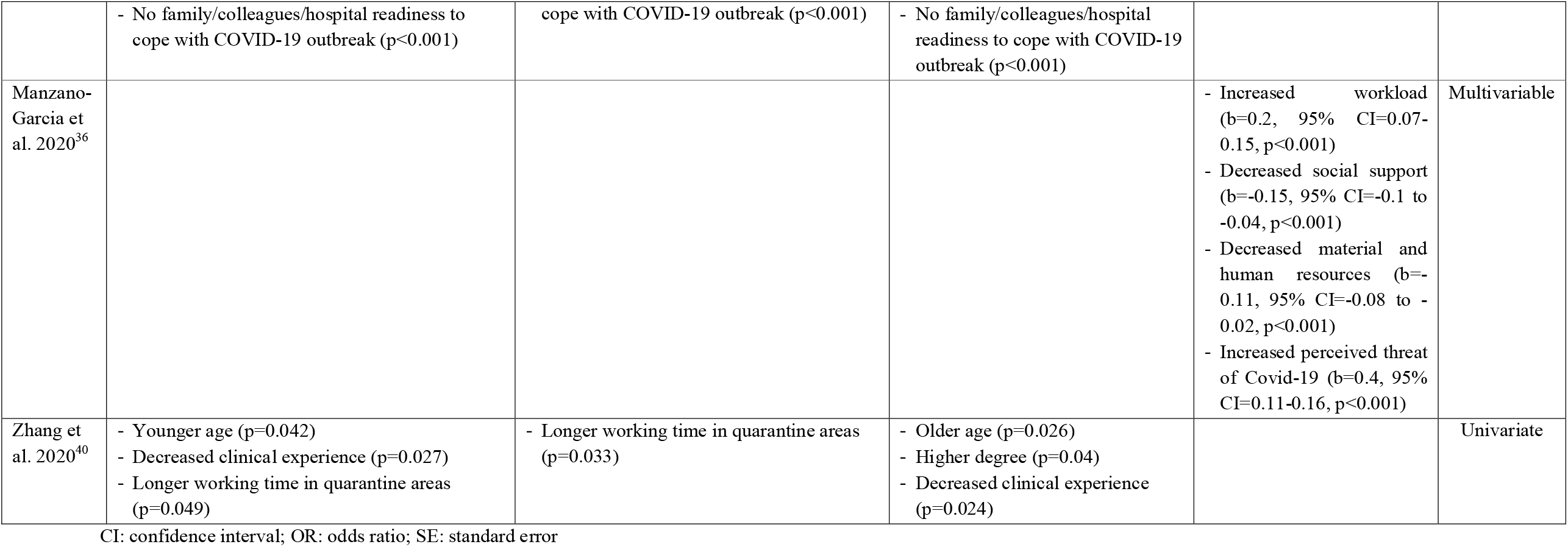
Risk factors for nurses’ burnout during the COVID-19 pandemic.

Sociodemographic factors (gender, age, educational level, and degree) affected nurses’ burnout. In particular, two studies^27,28^ found that females had higher levels of emotional exhaustion, but males had higher level of depersonalization^33^ and lower level of personal accomplishment^27^. Also, younger age,^28,40^ higher educational level,^28,33^ and higher degree^40^ increased nurses’ burnout.

Several social factors increased nurses’ burnout such as decreased social support,^36^ having a relative/friend diagnosed with COVID-19,^28^ low family and colleagues readiness to cope with COVID-19 outbreak,^33^ increased perceived threat of Covid-19,^36^ and longer working time in quarantine areas^40^.

Occupational factors affected nurses’ burnout during the COVID-19 pandemic to a large extent. Nurses that work in a high-risk environment e.g. a COVID-19 designated hospital, a COVID-19 unit or a critical care unit^27,27,28^ had higher level of burnout as well as nurses that work in hospitals with inadequate and insufficient material and human resources^36^. Also, nurses with lower levels of specialized training regarding COVID-19, job experience, and self-confidence in caring for COVID-19 patients experienced burnout more frequent^33,40^. Increased workload, decreased self-confidence in self-protection and decreased working safety while caring for COVID-19 patients were associated with increased burnout^33,36,40^.

## Discussion

We conducted a comprehensive systematic review to investigate the prevalence of nurses’ burnout during the COVID-19 pandemic and to identify associated risk factors. Also, we performed a meta-analysis examining the prevalence of three aspects of nurses’ burnout according to the MBI; emotional exhaustion, depersonalization, and lack of personal accomplishment.

We found a significant prevalence of nurses’ burnout during the COVID-19 pandemic according to the MBI. In particular, 34.1%, 15.2% and 12.6% of nurses experienced high levels of emotional exhaustion, low personal accomplishment, and depersonalization respectively. These levels of burnout are higher by far even among nurses working in a highly stressful environment such as palliative care; the prevalence of emotional exhaustion, low personal accomplishment, and depersonalization is 19.5%, 9.3%, 8.2% respectively^46^. Another meta-analysis^47^ included data from 49 countries and found that the overall prevalence of burnout symptoms among nurses is 11.23%. Nurses during the COVID-19 pandemic have higher level of emotional exhaustion, but lower level of depersonalization and higher level of personal accomplishment than mental health nurses,^48^ nurses in primary health care services,^49^ nurses in gynecology and obstetrics services,^50^ paediatric nurses,^51^ and emergency nurses^52^. Nurses’ daily emotions have been greatly challenged during the COVID-19 pandemic since they are a high-risk group, they have close contact with COVID-19 patients, and they are afraid of the consequences of the disease. The negative emotions and feelings of patients, colleagues, and family members can trigger similar emotions and feelings in nurses influencing perceived stress among them and making them more vulnerable to emotional exhaustion^53^. Also, higher job demands, workload, job complexity, job pressure, and working time during the COVID-19 pandemic increases work related stress among nurses resulting in emotional exhaustion. On the other hand, nurses during the COVID-19 pandemic experienced depersonalization and low personal accomplishment but not in higher levels than nurses working in stressful environment such as mental health nurses, nurses in primary health care services, nurses in gynecology and obstetrics services, paediatric nurses, and emergency nurses. This might be explained due to empathy and feelings that nurses have developed toward COVID-19 patients during a frightening situation that impacts all individuals’ lives such a pandemic. A pandemic may trigger compassionate behaviors among nurses connecting them with patients in a deeper level. Also, the effective treatment and care of COVID-19 patients improves nurses’ moral feeling competent and successful in their duty.

Apart from burnout, HCWs experience several other psychological and mental health outcomes during the COVID-19 pandemic such as depression, anxiety, post-traumatic stress disorder, psychological distress, sleep disturbances, insomnia, and fear^8–20^. The situation is even worse for nurses since it is well known that they struggle with burnout symptoms and other psychological issues more often than other HCWs resulting in negative consequences for themselves, their patients, their family members, their colleagues, and the health care organizations^12,46^. Nurses who are exposed and in contact with verified or suspected COVID-19 patients are more often distressed, nervous and frightened^54^. Moreover, nurses have extra concerns in COVID-19 pandemic such as the shortage of personal protective equipment, and the fear of being exposed at work, spreading the SARS-CoV-2, and taking the virus home to their close family members^9^. Thus, nurses are facing a continuous stress that can trigger post-traumatic stress, suicide ideation and suicide^55^. Also, this stress results in burnout that can negatively affect the quality of health care that nurses provide to patients^56^.

We found that several sociodemographic, social and occupational factors increase nurses’ burnout during COVID-19 burnout. Gender is a controversial issue since our review showed that females have higher levels of emotional exhaustion, but males have higher level of depersonalization and lack of personal accomplishment. After exposure to stressful events, females were more likely to be traumatized than males^57–59^. In contrast, a meta-analysis^60^ with 57 studies found that being male is related to higher level of burnout among nurses. Moderator variables (age, job satisfaction, position, clinical experience, etc.) should be taken into consideration to infer more valid results regarding the role of gender on nurses’ burnout. Younger nurses are more likely to exhibit burnout during COVID-19 pandemic than older nurses and this might be related to the fact that younger are less familiar with infection control and protective measures and less experienced in handling extreme events such as a pandemic^61^. Probably, younger nurses are more vulnerable when facing difficult situations such as patients suffer and die from COVID-19 especially in cases where HCWs cannot offer the standard health care due to sources limitations.

According to our review, decreased social support is associated with increased nurses’ burnout during COVID-19 pandemic. Psychological support that HCWs receive during and after a pandemic can significantly influence their feelings and emotions handling in a better way the negative effects of such a devastated event^62,63^. Also, we found that longer working time in quarantine increases nurses’ burnout. This result is confirmed by studies with nurses that work in quarantine areas during epidemics where loneliness has been recognized as a major stressor^64,65^. Loneliness is magnified in cases that nurses have to separate from their families and stay at designated hospitals as has happened in Wuhan, China^40^. Especially family and social support is an essential weapon for nurses to confront the psychological distress that experience during epidemic outbreaks^66^. A systematic review found that lack of social support is an important risk factor for the development of psychological issues in HCWs during disasters^67^. Support from families, friends, colleagues, and health care organizations gives nurses the opportunity to control effectively and avoid negative feelings and emotions decreasing the risk of burnout syndrome. In particular, several studies show that a strong social support network during the COVID-19 pandemic can decrease feelings of isolation and strength resilience among HCWs^68–70^.

Moreover, we found that nurses having a relative/friend diagnosed with COVID-19 experience a higher level of burnout. The COVID-19 pandemic and public health response to it ultimately had changed work and life conditions such as other epidemic outbreaks e.g. SARS. In that case, nurses worry more about the health of their close family members/friends/colleagues than their own^61,71^. Nurses try to avoid close contact in purpose, to reduce the spread of SARS-CoV-2 to their family members/friends/colleagues. Thus, nurses’ home and social life is significantly disrupted resulting in fear, anxiety, and psychological distress. Also, caring for relatives/friends/colleagues as patients is emotionally difficult and exhausted. Especially in case that nurses take care of their colleagues suffer from the fear of their own personal vulnerability^61,71^.

We found that nurses working in a high-risk clinical environment (a COVID-19 designated hospital, a COVID-19 unit, hospitals with inadequate and insufficient material and human resources, low working safety while caring for COVID-19 patients, etc.) have higher level of burnout. This finding is confirmed by previous research during the SARS outbreak^72^. A high-risk clinical environment is an important source of distress for nurses increasing feelings of loss of control or vulnerability and concerns about spread of SARS-CoV-2, health of family members/friends/colleagues, and changes in home and work life^73^. In addition, severe shortage in personal protective equipment and nursing staff and the increasing number of suspected and actual COVID-19 cases add more pressure to nurses^74^.

Our review identified that poor working conditions such as increased workload, low level of specialized training regarding COVID-19, and increased working times increase level of burnout among nurses. Several studies have already shown that nurses exhibit burnout due to the prolonged direct personal contact with a great number of patients^75,76^ as well as the inadequate staffing and resources^77–79^. Also, poor working conditions are an important risk factor for work-related stress and job dissatisfaction that end up on high levels of emotional exhaustion, depersonalization, and low level of personal accomplishment among nurses^80–83^. Low level of specialized training regarding COVID-19 is an issue that needs special attention since knowledge, control measures and personal protective equipment against COVID-19 are limited. Pandemic (H1N1) 2009^84^ and SARS epidemic^85^ underline nurses’ concern about inadequate training and expertise in handling challenging health care issues. There is a need for nurses to obtain new knowledge and skills about COVID-19 to built their confidence in providing health care under this extreme situation. Well-trained nurses could improve their self-efficacy that is necessary to confront disasters such a pandemic^86^.

There were some limitations in this systematic review and meta-analysis. First, the majority of studies were of moderate quality, while only five studies included entirely nurses. Also, due to the limited data and the limited number of studies, subgroup analysis and meta-regression analysis cannot be performed. Data regarding risk factors for nurses’ burnout during the COVID-19 pandemic were available only in five studies and among them three used multivariable methods to eliminate confounders. Second, we searched three major databases and pre-print services as well as the full reference lists of all selected articles but there is still the probability to not identify some studies e.g. in grey literature. Third, the heterogeneity between results was very high and we applied random effects model to handle with this issue. Forth, we included studies that used standardized and valid instruments to measure burnout but these data are still self-reported and inherently subjective. Moreover, only cross-sectional studies with convenience or snowball sampling method were identified making definite causal relationships impossible.

In conclusion, nurses experience high levels of burnout during the COVID-19 pandemic, while several sociodemographic, social and occupational factors affect this burnout. The COVID-19 pandemic is a significant challenge for nurses worldwide and learning lessons from the first wave is imperative to prepare better strategies for the subsequent waves. Several measures could be introduced to mitigate mental health impact of the COVID-19 pandemic on nurses, e.g. screening for mental health illness and early supportive interventions for high-risk nurses, immediate access to mental health care services, designated rest periods, social support through hospital support groups to reduce feelings of isolation, sufficient personal protective equipment for all nurses to provide security etc. As the second wave of the COVID-19 pandemic is hitting worldwide and there are predictions for following waves in the near future, there is a need to decrease nurses’ burnout and improve their mental health. Governments, health care organizations and policy makers should act in this direction to prepare health care systems, individuals and nurses for a better response against the COVID-19 pandemic.

## Supporting information

Web Table 1. PRISMA Checklist

Web Figure 1. A leave-one-out sensitivity analysis of the prevalence of emotional exhaustion among nurses according to the Maslach Burnout Inventory.

Web Figure 2. Funnel plot of the prevalence of emotional exhaustion among nurses according to the Maslach Burnout Inventory

Web Figure 3. A leave-one-out sensitivity analysis of the prevalence of depersonalization among nurses according to the Maslach Burnout Inventory

Web Figure 4. Funnel plot of the prevalence of depersonalization among nurses according to the Maslach Burnout Inventory

Web Figure 5. A leave-one-out sensitivity analysis of the prevalence of lack of personal accomplishment among nurses according to the Maslach Burnout

Web Figure 6. Funnel plot of the prevalence of lack of personal accomplishment among nurses according to the Maslach Burnout Inventory

## Data Availability

Data will be available after request.

## Conflicts of interest

none

## Funding

None

## Declarations of interest

None

**Web Figure 1**. A leave-one-out sensitivity analysis of the prevalence of emotional exhaustion among nurses according to the Maslach Burnout Inventory.

**Web Figure 2**. Funnel plot of the prevalence of emotional exhaustion among nurses according to the Maslach Burnout Inventory.

**Web Figure 3**. A leave-one-out sensitivity analysis of the prevalence of depersonalization among nurses according to the Maslach Burnout Inventory.

**Web Figure 4**. Funnel plot of the prevalence of depersonalization among nurses according to the Maslach Burnout Inventory.

**Web Figure 5**. A leave-one-out sensitivity analysis of the prevalence of lack of personal accomplishment among nurses according to the Maslach Burnout Inventory.

**Web Figure 6**. Funnel plot of the prevalence of lack of personal accomplishment among nurses according to the Maslach Burnout Inventory.

## Notes

### Competing Interest Statement

The authors have declared no competing interest.

### Author Declarations

There is no need for approval of the IRB/oversight body

