## Supplementary figures and images for "Nurses’ burnout and associated risk factors during the COVID-19 pandemic: a systematic review and meta-analysis"

### Web Figure 1. A leave-one-out sensitivity analysis of the prevalence of emotional exhaustion among nurses according to the Maslach Burnout Inventory.

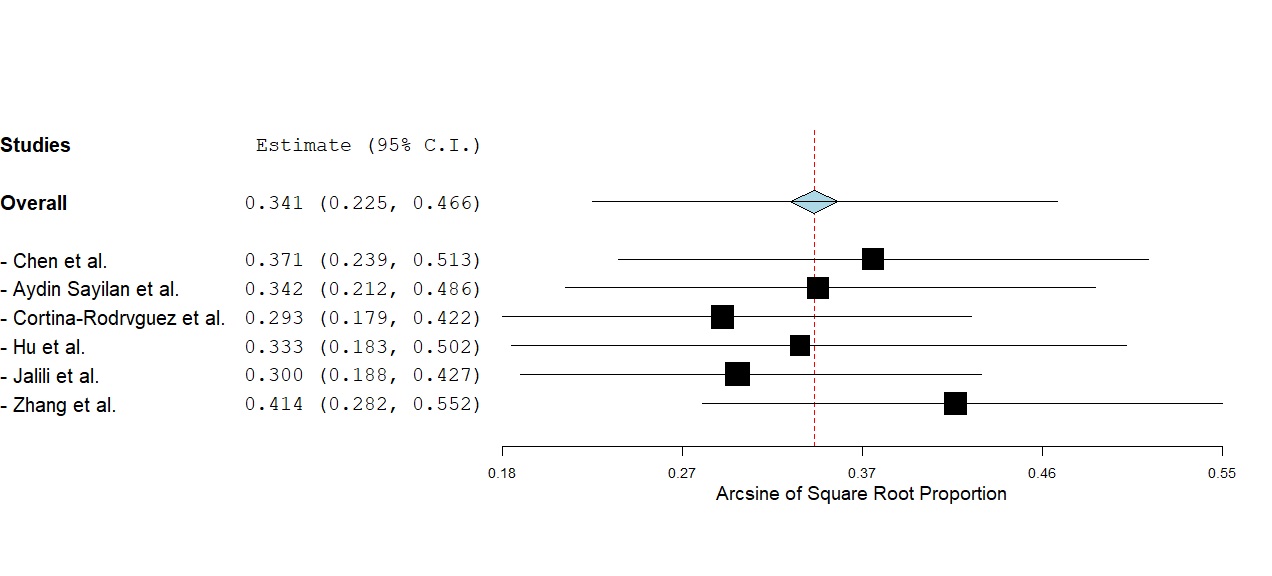

### Web Figure 2. Funnel plot of the prevalence of emotional exhaustion among nurses according to the Maslach Burnout Inventory

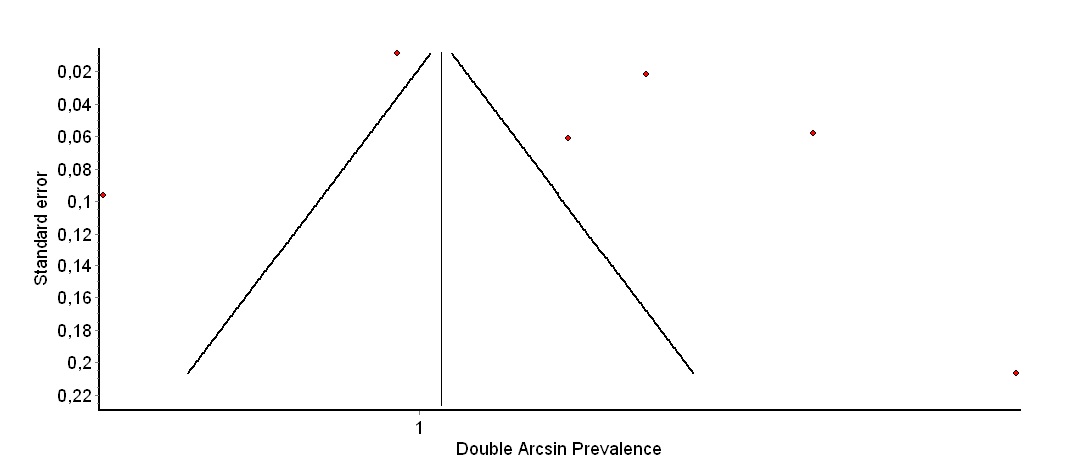

### Web Figure 3. A leave-one-out sensitivity analysis of the prevalence of depersonalization among nurses according to the Maslach Burnout Inventory

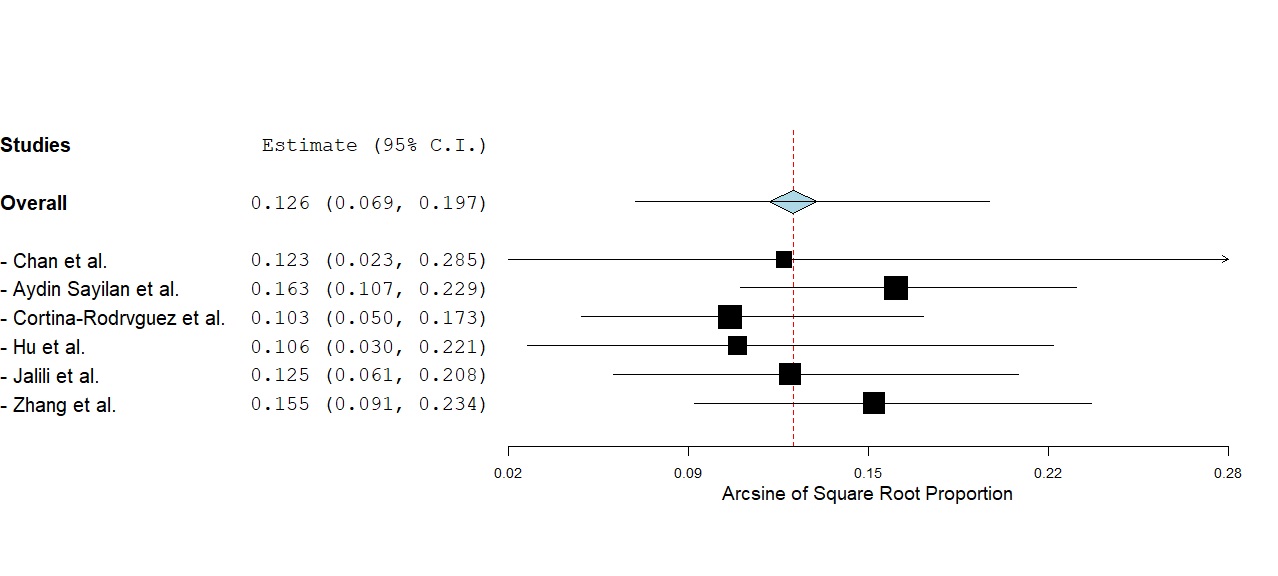

### Web Figure 4. Funnel plot of the prevalence of depersonalization among nurses according to the Maslach Burnout Inventory

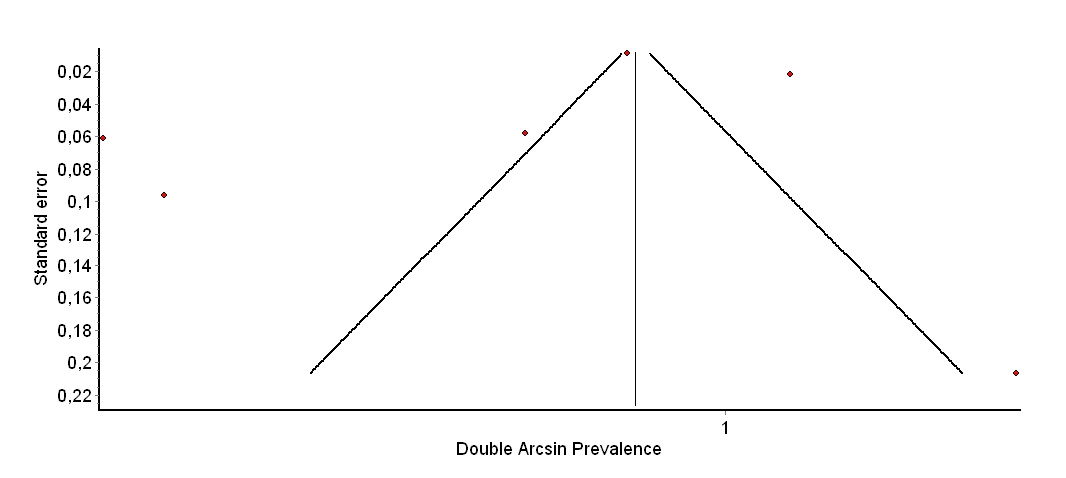

### Web Figure 5. A leave-one-out sensitivity analysis of the prevalence of lack of personal accomplishment among nurses according to the Maslach Burnout

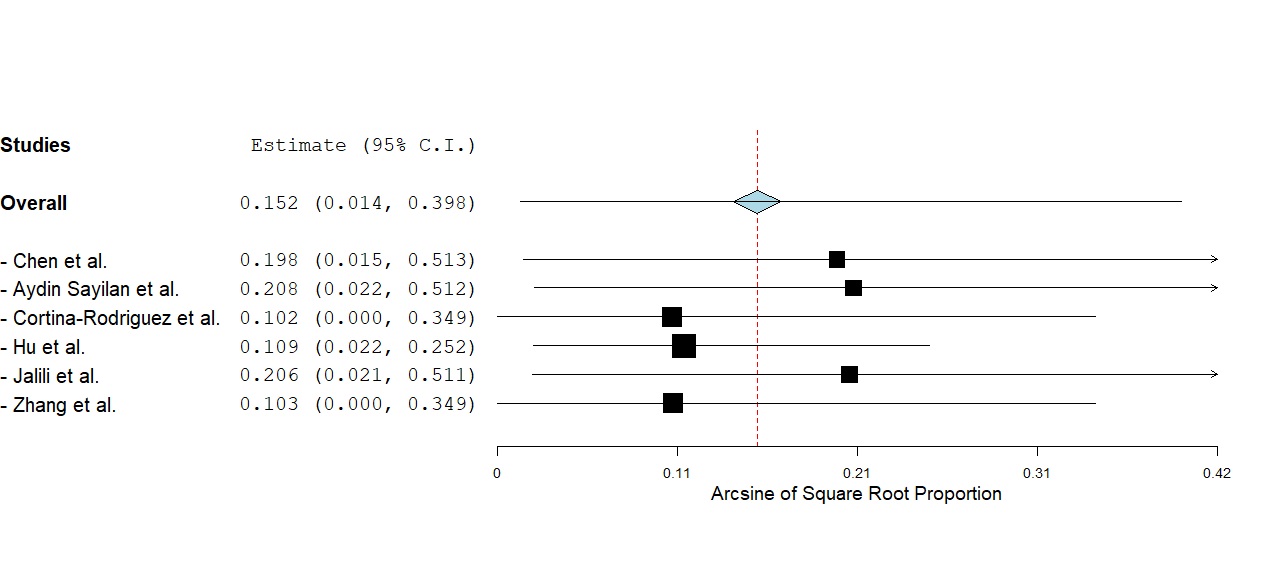

### Web Figure 6. Funnel plot of the prevalence of lack of personal accomplishment among nurses according to the Maslach Burnout Inventory

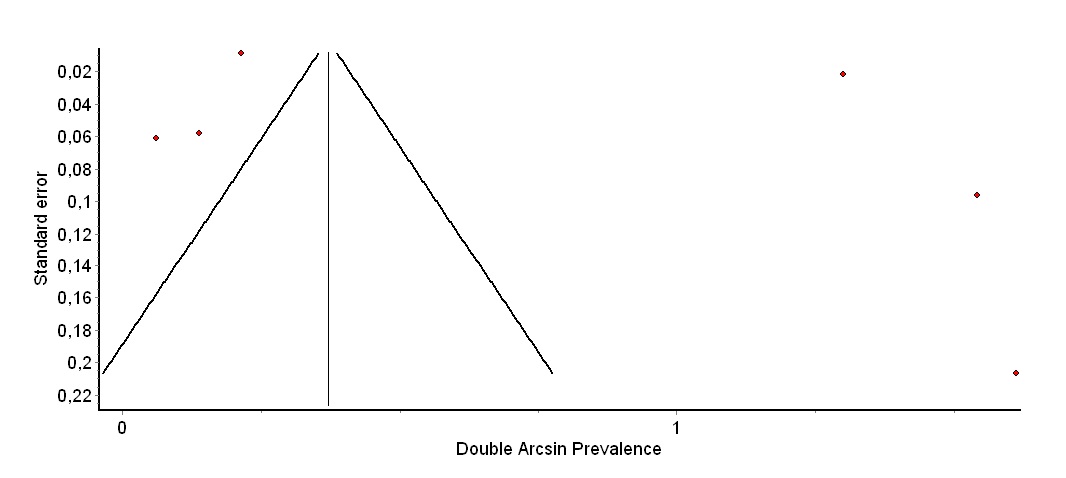
